# A causally informed framework for robust confounder control in biomedical machine learning

**DOI:** 10.1101/2024.09.20.24314055

**Authors:** Vera Komeyer, Carolin Herrmann, Simon B. Eickhoff, Charles Rathkopf, Federico Raimondo, Kaustubh R. Patil

## Abstract

Machine learning (ML) offers transformative opportunities for neurobiomedicine, yet predictive models often exploit confounding-driven associations rather than genuine biological mechanisms, undermining generalizability and neurobiomedical validity. Current practice commonly defines confounders heuristically (e.g., age, sex) or correlationally, risking confusion with colliders or mediators. To address this, we propose a pragmatically integratable, causally informed three-step framework for confounder selection and adjustment aimed to support debiased, meaningful neurobiomedical supervised ML (SML) models. Step 1 involves a domain-knowledge-driven causal analysis of a specific research question, formalized in a directed acyclic graph (DAG). Step 2 applies graph-theoretic rules to the DAG to identify valid deconfounding variables. Additionally, it provides strategies for unmeasured variables, including discussion of their theoretical and practical strengths and limitations. Step 3 integrates the causal justification with empirical associations, ensuring that only statistically relevant confounders are adjusted for. We illustrate the framework’s practical application using a UK Biobank-based brain-behaviour prediction example and demonstrate the substantial impact of confounding on predictive models – underscoring the necessity of proper deconfounding. Despite the populartity of linear feature residualization, its reliance on linear assumptions and adjustment of only features (or target) limits its effectiveness. As a potential solution, we introduce double machine learning, originally developped for causal inference, and discuss its adaptability to associative SML. Importantly, causally informed deconfounded SML models should not be causally interpreted without further justifications. Nevertheless, they are essential for producing robust, generalizable, and neurobiomedically meaningful predictive insights.

## 1. Introduction

### 1.1. Predictive analytics are a useful tool in neurobiomedicine if models are unbiased

Machine learning (ML) and artificial intelligence (AI) offer transformative potential in neurobiomedical research and deployment. Using large, high-dimensional and oftentimes observational datasets, ML enables the development of predictive models for identifying biomarkers and supporting diagnosis, prognosis and treatment decisions ^1–3^. Predictive models thereby serve two main purposes: (1) supporting scientific discovery by uncovering neurobiological mechansims and (2) advancing precision medicine through clinical decision-making tools (e.g. ^4^).

Both goals require reliable models that generalize across settings. Scientific models (case 1) aim to answer scientific questions such as identifying brain patterns linked to psychiatric conditions (e,g. depression or schizophrenia ^5,6^). Achieving such a deeper understanding of neurobiomedical mechanisms requires reliable and generalisable insights. Clinical (tools) models (case 2) must provide generalizable predictions for consistent usability across hospitals and patient populations, akin to standardized diagnostic tests. However, in both cases, the conventional emphasis on merely maximizing model accuracy, can cause models to fail when applied to new conditions ^7,8^, e.g. due to data distribution shifts ^9^ or covariate shifts ^10,11^ if the high accuracy was based on models overfitting to specific datasets. Problematically, such failure of generalization is often accompanied by unreliability of predictions ^12–15^. A sole focus on accuracy maximisation hence risks overlooking of biological and clinical meaningfulness of models.

One important but often underappreciated source of poor generalization and unreliable predictions – beyond issues such as small sample sizes, poor regularization or model misspecification-is bias. Biased models base their predictions on spurious associations rather than genuine biological relationships (**Figure 1**a), hindering generalizability to new data. A key contributor to biased supervised machine learning (SML) models is confounding, where a variable influences both the input features and target outcomes (**Box 1**; confounder bias, Simpson’s Paradox). For example, in psychiatric research, comorbid conditions (e.g. anxiety, substance use disorder), age or medication use can influence both brain imaging features and psychiatric diagnoses, introducing misleading associations and making it difficult to disentangle disorder-specific neural signatures from overlapping, yet distinct, effects. For instance, a model might falsely attribute structural brain changes to schizophrenia when, these changes are actually (partially) driven by aging or long-term medication use. In essence, biased models incorrectly attribute effects such as a psychiatric condition to features (e.g. brain measures) while the effects are actually due to another variable (the confounder), which limits both generalizability and biological insights of a model.

**Figure 1.**
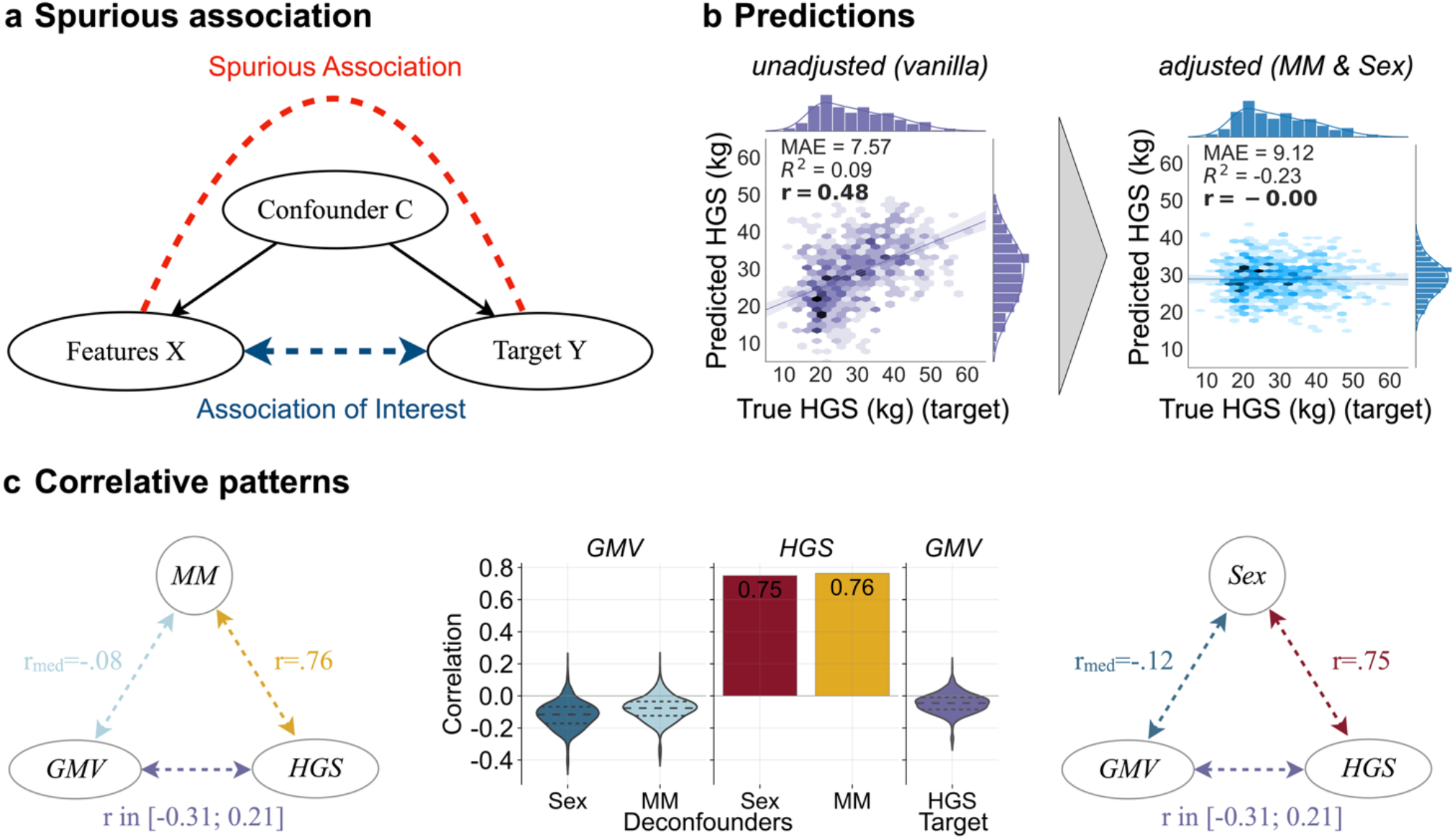
Illustration of concepts and statistical relationships of variables involved in the illustrative prediction of hand grip strength (HGS) from gray matter volume (GMV). **a.** When investigation the relationships between features X and a target Y, a confounding variable C can introduce a spurious association, biasing the actual association of interest as it influences both the features X and the target Y (inspired by ^26^). **b**. Prediction of HGS from 1088 parcellated cortical, subcortical and cerebellar GMV features in N=3620 healthy subjects from the UK Biobank using a linear support vector regression (SVR) not adjusted for confounding (left) and adjusted for the identified deconfounders muscle mass (MM) and sex (linear feature regression) (right). **c**. Pairwise statistical association between features (GMV), target (HGS) and both deconfounders (MM, sex). GMV refers to a feature vector with 1088 features so that correlations with the deconfounders are indicated as median value (left and right plot, blue arrows) and correlation with the target as range (purple arrows). Violin plots (middle plot) show the respective distributions of parcel-wise feature associations with the deconfounders (blue) and the target (purple).

Complex variable interdependencies in neurobiological research make it challenging to identify which third variables (those that are neither features nor targets) qualify as confounders (and which do not). In many biomedical disciplines it is common to correct for a conventionally established set of confounders (e.g. demographics, lifestyle factors etc.), without transparent and systematic justification ^16–18^. When justifications are provided, they often rest solely on report of statistical associations between potential confounders and feature(s) (X) and/or target (Y) ^18–20^. However, neither no justification nor justification through correlative patterns is enough. Instead, effective confounder identification requires understanding the causal roles of third variables in the context of a specific research question.

### 1.2. Causal justification is required for proper deconfounding

Correlation-based definitions alone are not enough because different types of third variables Z, namely confounders, colliders and mediators **(Box 1)** could produce the same correlation between Z and both X and Y ^18^. This is problematic because only confounders should be controlled for, while making sure to not correct for colliders as this would inadvertently introduce bias (collider bias (e.g. ^21,22^), Berksons’s paradox ^23^, **Box 1**). This implies that adjusting for a third variable is not always right or not adjusting for it is always wrong. Rather, the decision depends on the process that generated the data, which is what we are ultimately interested in to achieve both, better transportability and generalizability of models and to better understand biological mechanisms. Consequently, arbitrary or standard adjustments risk introducing rather than removing bias. While correlations do not differ between types of third variables, directionalities i.e. causalities do differ (**Box 1**).

Directed acyclic graphs (DAG, **Box 2**) offer a principled way to clarify variable roles and systematize relationships by encoding causal assumptions (directionalities) around the relationship of interest. Additionally, they enable transparent communication of assumptions and are hence critical for building unbiased SML models ^24,25^. Constructing such DAGs requires domain expertise and literature-based causal reasoning. As first objective, here, we will illustrate, how building such a DAG can be achieved in the context of biological SML tasks, by leveraging a typically applied bottom-up strategy from classical statistical causal inference to foster unbiased meaningful model development.

### 1.3 Challenge of unmeasured confounders in neurobiomedical observational data

Even with a principled, DAG-informed approach to confounder selection, a critical challenge remains: relevant confounders may be unmeasured or entirely unobservable. This is especially common in neurobiomedical contexts, where important biological constructs (e.g. hormone levels, early-life adversity, genetic liabilities) may be latent, unrecorded, or infeasible to measure. In such cases, standard confounder selection strategies, such as the so-called backdoor adjustment (see chapter 2, step 2 and **Box 3**), fall short, and alternative strategies are necessary. To tackle this issue, here as second objective we will introduce and discuss how a range of established methods from the causal inference literature can be applied to the SML context to handle unmeasured confounding. We neither claim novelty nor completeness of presented methods, but aim to build bridges between disciplines by suggesting and discussing integrability and feasibility of tools for debiasing of neurobiomedical SML models.

In summary, unaddressed confounding or improper handling of other third variables can lead to biased SML models based on spurious associations, limiting their generalizability and interpretability issues that undermine model utility for clinical applications and mechanistic insights. Correlation-based confounder selection, though commonly used, is inadequate. Because confounding is inherently causal, we here advocate for the integration of causal reasoning into the confounder selection process for SML workflows. Importantly, our goal is not to estimate treatment-outcome effects, generate counterfactual data or build causal discovery graphs (identifying the correct DAG from data) such as pursuit in traditional causal inference frameworks and causal ML approaches. Rather, we aim to make causal inference principles accessible and practically useful for researchers in neurobiomedical SML. Concretely, we propose an easy-to-follow stepwise framework for identification of a suitable set of third variables to adjust for, and offer practical guidance for unobserved or unmeasured confounders (chapter 2). Additionally, we discuss the limitations of post-hoc linear (feature) residualization and explore the potential of adopting Double Machine Learning (DML) as an alternative confounder adjustment strategy (chapter 3). While deconfounded ML can support unbiased models, it does not equate to causal inference (chapter 4). By making causal inference principles accessible to researchers working in neurobiomedical SML we aim to link statistical prediction with causal reasoning. This offers a structured way to remove bias so that confounder adjustment enhances rather than hinders model validity, reliability and generalizability.

## 2. 3-step framework for confounder selection and adjustment

The core mechanism for unbiasing supervised ML models is through the identification of and adjustment for a correct set of deconfounders. While confounders are all variables that confound the X-Y relationship, deconfounders are a sufficient subset of confounders whose adjustment blocks non-causal (confounding) paths between input features (X) and the target (Y) and enables unbiased prediction (**Box 3**). Identification of a correct set of deconfounders requires a causal analysis around the relationship of interest (X-Y) to identify different possibilities for confounder adjustment (**Figure 2**, step 1). Additionally, unbiasing may require strategies to handle cases of unobserved confounders (once identified), an ubiquitous problem when using observational data as often the case in neurobiomedical supervised ML (**Figure 2**, step 2). Once all adjustment variables have been identified the statistical relevance of variables must be confirmed before adjusting the supervised ML model (**Figure 2**, step 3). In the following we will detail each step and discuss benefits and challenges in the context of deconfounding a supervised ML model. The fundament for all steps is built by a causal analysis which is summarised in a directed acyclic graph (DAG) (**Box 2**). As this analysis relies on domain knowledge about the process that generates the observational data all steps are research-question dependent, i.e. there is no one-size-fits-all solution. We therefore exemplify each theoretical step with a real-world example of a supervised prediction.

**Figure 2.**
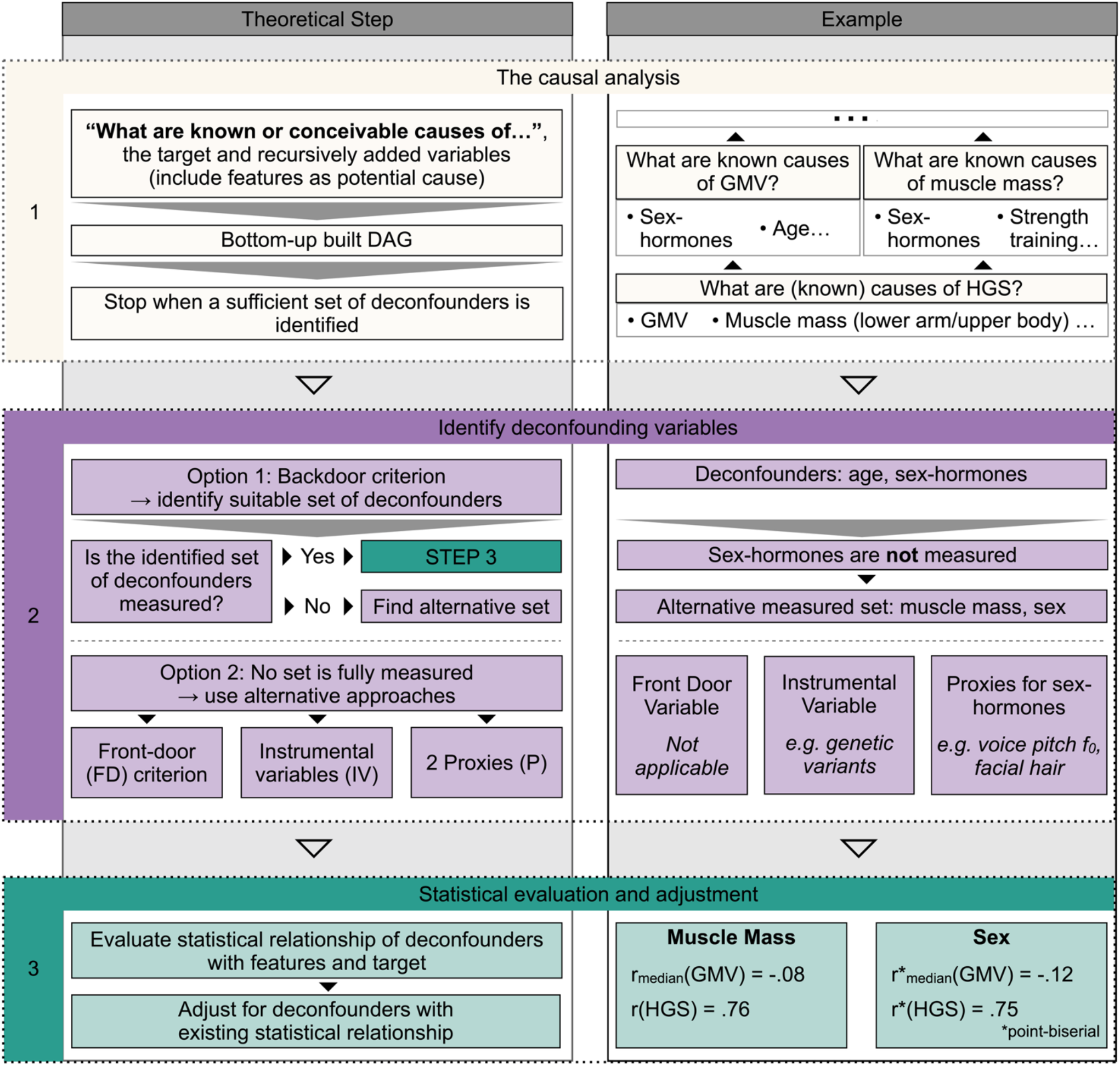
3-step framework for confounder selection with left panel describing the theoretical step and the right panel illustrating it using the example of predicting hand grip strength (HGS) from parcellated gray matter volume (GMV). **Step 1** (beige) involves the causal analysis around the research question of interest, illustrating bottom-up building of the DAG. A sufficient set of deconfounders becomes clear from performing step 2 (chapter 2.2). **Step 2** (purple) involves different possibilities for identifying a proper set of deconfounding variables from the DAG built in step 1. The backdoor criterion is the easiest applicable approach (chapter 2.2.1), but in case of unmeasured deconfounders usage of the frontdoor criterium, instrumental variables or two proxies can serve as alternatives (chapter 2.2.2). **Step 3** covers the statistical evaluation of the identified deconfounders with the features and the target as well as the statistical adjustment of the model. In the example prediction, muscle mass was correlated with the 1088 GMV parcels in median by r_median_=-0.08 and with HGS by r=.76 and sex was correlated (point-biserial correlation) with GMV by r_median_=-0.12 and with HGS by r=.75 (**Figure 1**).

Concretely, the SML example is the out of sample prediction of the target Hand Grip Strength (HGS) from T1w-MRI derived Grey Matter Volume (GMV) features in the UK Biobank (UKB) ^27^ as a large observational dataset (for methods see supplementary materials). In this example, a simple linear support vector regression (SVR) prediction model (see methods) without confounder consideration (*vanilla* model) can lead to a decent prediction as indicated by a correlation between true and predicted HGS of r=0.48 (**Figure 1b**, left). However, the following outlined stepwise approach for correct deconfounding will clarify that this model is biased, i.e. the decent prediction is biased by confounding signals.

### 2.1. Step 1 – The causal analysis

Conducting a causal analysis around the relationship of interest between input features (X) and the target variable (Y) is the cornerstone for all subsequent steps. This analysis is formalized using a DAG, which serves as a structured representation of the assumed causal relationships among all relevant variables. The DAG allows to differentiate between various types of third variables (e.g. confounders, mediators, colliders) and thereby helps to identify a suitable set of variables to adjust for to block confounding paths between X and Y.

We here suggest a bottom-up strategy to guide the causal analysis and determine influential factors on X and Y. This begins by asking about known and conceivable causes of the target Y. Starting with Y, additional variables are iteratively added based on their potential causal influence – either on Y or on other already included variables, until the network of relevant relationships is mapped. Constructing valid directed edges (arrows) in the DAG requires domain knowledge and literature justification to encode both empirically established and theoretically plausible cause-effect relationships ^28^. While the process is inherently subjective, its strength lies in making modelling assumptions explicit and transparent, in contrast to arbitrary or purely correlation-based confounder selection. In the GMV-HGS example, the DAG might start with *lower arm/upper body muscle mass* as established physiological cause of HGS (muscle mass → HGS) and sex-specific influences on HGS independent of muscle-mass e.g. through sex differences in muscle function. Additionally, the GMV is added as conceivable cause of HGS as GMV-HGS is the hypothesized predictive relationship to be unbiased^1^. In the next iteration, known or conceivable causes of *muscle mass* could be *sex hormones, eating behaviour, strength training, age* etc.. The GMV features are influenced by *TIV*, age, *sex hormones* and further -potentially unmeasurable or unobserved - environmental and behavioural factors. Iterating this process builds a DAG (**Figure 3**), where the bottom-up approach allows to systematically disentangle the complex causal structure of intertwined biological, environmental, and behavioural factors typical for neurobiomedical data.

**Figure 3.**
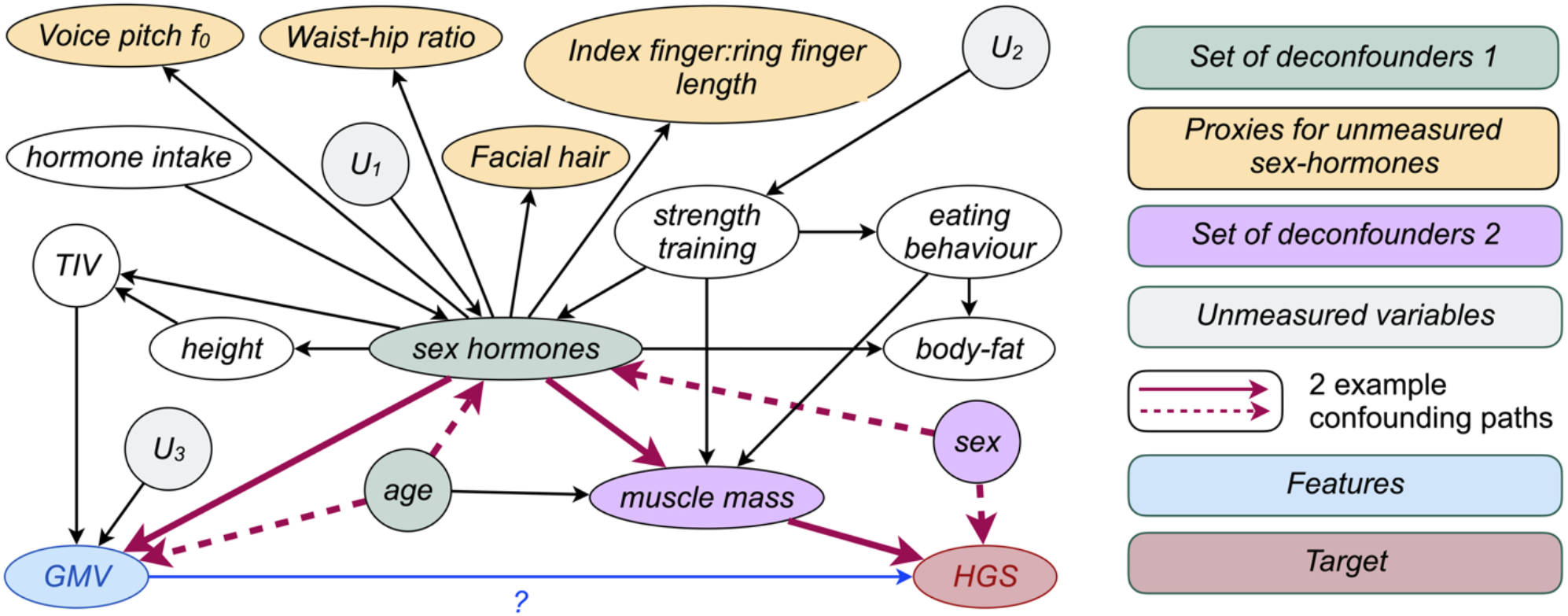
Example DAG for deconfounder identification for the GMV-HGS prediction example resulting from causal analysis following the framework outlined in Figure 2. Two example confounding paths are highlighted in red (not comprehensive). Considering all confounding pathways, one potential minimal set of adjustment variables (deconfounders) is *sex hormones* and *age* (set of deconfounders 1). As *sex hormones* are unmeasured in the exemplary data sample, an alternative sufficient set of deconfounders would be *muscle mass* and *sex* (purple). Both sets qualify to block all non-causal pathways following the backdoor criterion and are hence a sufficient subset of variables to debias the predictive GMV-HGS model.

A key challenge in confounder selection for predictive modelling is knowing when and if all confounders were identified. This is where the concept of deconfounders becomes useful: Rather than attempting to include *all* confounders, the goal is to adjust for a sufficient subset that blocks all so-called backdoor paths between X and Y (see **Box 3** and step 2). The bottom-up construction process helps determine when adding further variables does not yield meaningful gains in bias reduction. For example, further specification of an unmeasured variable *U*_*2*_ in **Figure 3** would not change the deconfounding status of the GMV-HGS relationship. In step 2 we will discuss existing options to identify a sufficient set of deconfounders.

While the DAG relies on established causal relations, oftentimes not all links are well-known (yet). In cases where domain knowledge is incomplete or even contradictory, the DAG must rely on ambiguous cause-effect assumptions, so that multiple plausible DAGs may exist for the same research question. Importantly, causal assumptions encoded in a DAG cannot be empirically verified using observational data alone, and the bias from incorrect assumptions does not vanish with large sample sizes ^29^. To address this well-known challenge, the field of causal graph discovery offers methods for learning DAGs from data (e.g. DAG-GNN, CAM, NOTEARS, LiNGAM, GES etc.), some of which have been applied to uncover causal structures in areas such as Alzheimer’s disease biomarkers ^30^. However, such approaches are often (computationally) comprehensive research projects and therefore not feasible as part of another project that targets the out-of-sample prediction of a feature-target relationship rather than modelling the causal assumptions around this relationship.

Despite its limitations, a hypothesis- and literature-based DAG remains a powerful tool. Through clear justification it enables formalization and transparent communication of assumptions, strengthening replicability of the causal reasoning underpinning a model. Additionally, the DAG provides a basis for interpreting model outputs contingent upon the used set of deconfounding variables, based on the assumptions the DAG advertises. Perhaps most importantly, the causal analysis forces researchers to precisely *think* about and critically engage with the question the model is about to answer - an aspect sometimes falling short in pure data-driven approaches.

### 2.2. Step 2 – Identifying a suitable set of deconfounders and options in the case of unobserved deconfounders

#### 2.2.1. Identifying a suitable set of deconfounders based on the backdoor criterion

Unlike purely correlative approaches, causal analysis as formalized in a DAG distinguishes confounding pathways from colliders and mediators (**Box 1**), enabling principled deconfounding strategies (**Box 3**). The most direct method for identifying a suitable set of deconfounders is based on the backdoor criterion ^25^. A set of variables Z satisfies the backdoor criterion relative to the relationship X-Y if (1) no variable in Z is a descendant of X, and (2) Z (all variables in the set) blocks all backdoor paths from X to Y (**Box 3**). Graphically, this means that variables with arrows pointing into X in the DAG qualify as valid adjustment variables Z, which when conditioned on, block non-causal pathways (flow of information) between X and Y. Such deconfounder sets can be identified manually by following graph rules in the DAG or automatized using dedicated tools (e.g. DAGitty ^28^ or CausalFusion (https://causalfusion.net)).

In the GMV-HGS example DAG (**Figure 3**), several confounding (backdoor) paths exist between GMV and HGS. One valid deconfounder set includes *sex-hormone levels* and *age*. Adjusting for this set would block all backdoor paths and hence debias the prediction of HGS from GMV (**Figure 3**, green). In practice, however, identified deconfounders may not be measured or unobservable. For example, in the GMV-HGS prediction, although *sex-hormone levels* are included in the UKB, their measurement occurred on average 14.79 years prior to the brain imaging (GMV) and HGS assessment (session 0 vs. session 2). Given this substantial temporal gap, using *sex-hormone levels* would not be valid for adjustments.

The first, and easiest way to overcome the problem of an unmeasured (set of) deconfounder(s) is by still applying the backdoor criterion but trying to find alternative sets of deconfounders (alternative routes) that would block all non-causal pathways. In the GMV-HGS example such an alternative set could be *sex* and *muscle-mass* (**Figure 3**, purple), both of which are measured in the UKB for session 2. If such alternatives exist (as in our example) one can proceed to step 3 of the process. However, there is a multitude of cases which lack any alternative measured deconfounder sets that satisfy the backdoor criterion. The next sections therefore elaborate on three alternative strategies: front-door adjustment, instrumental variables and proxies.

#### 2.2.2. Alternatives in the case of unobserved deconfounders

##### 2.2.2.1. Front door criterion

One alternative if deconfounders identified through the backdoor criterion are unavailable is front-door adjustment ^25^. It requires an intermediate variable F that meets three conditions: a) F intercepts all direct paths from X to Y (i.e., X → F → Y, b) there is no backdoor path from X to F and c) all backdoor paths from F to Y are blocked by X (**Box 3**). If these conditions are satisfied, the unbiased relationship between X and Y can be estimated by combining the estimate of effect X→F and of F→Y, circumventing the unobserved variable Z. In SML, this can be operationalised through two-stage models: the first model predicts F from X and the second predicts Y from predicted F.

In practice, however, finding such a variable F is especially challenging in neurobiomedical contexts. For example, in the GMV-HGS context, F would need to be causally affected exclusively by GMV and independently affect HGS, without being subject to the same confounding structure—this is typically hard to guarantee in neurobiomedical settings.

##### 2.2.2.2. Instrumental variables (IVs)

Another frequently used alternative in the presence of unmeasured confounders are instrumental variables (IVs) ^31^. A third variable V qualifies as IV if it satisfies three assumptions: (a) independence: The unmeasured confounder Z and V are independent (no arrow) (in short V⊥Z); (b) relevance: V causes X (V→X); (c) exclusion restriction: V affects Y only through X (V→X→Y) (no direct causal connection V→Y) (**Box 1**). Conceptually, IVs can be understood as mimicking randomization or simulating an experimental intervention. They thereby provide variation in X that is orthogonal (independent) to the confounding structure. As there are no confounders of the relation between V and Y, any observed association must be causal. Moreover, since V affects Y only via X, V allows to isolate the variation in X free of confounding, enabling causal estimation even when Z is unobserved. In SML practice, IVs are used in a two-stage process, where in the first stage V is used to predict X and this predicted X is used to predict Y. For example, in our neurobiological application, genetic variants (SNPs) identified in genome-wide association studies (GWAS) could serve as potential IV if there were a genetic variant known to affect GMV but assumed to not directly affect HGS.

While standard IV, as applied for causal inference, assumes linear relationships between V and X (e.g. modelling the first stage as X=πV+ϵ), the particular strength of transporting this approach to the SML context is that using any kind of ML model for the first stage prediction can better capture complex, nonlinear and interactive relationships between V an X. This is especially suitable for the oftentimes high-dimensional setup of SML with potentially complex instruments and features. Therefore, ultimately, the usefulness of this approach relies on the strength of the IV V and on how well the features X are predicted from V: the better V and the better the prediction of X, the better and less biased the final estimation of Y (in which we are interested) is.

However, IV estimation comes with trade-offs. Most notably, it can exhibit higher variance compared to direct confounder adjustment methods, especially when the IV is weak (i.e. V only weakly predicts X). Weak instruments introduce noise in the first-stage prediction, leading to noisy and unstable second-stage estimates ^29^. This resembles a classic bias-variance trade-off: while strong and valid IVs can eliminate systematic bias from unmeasured confounding, especially weak IVs can lead to high variance estimates. This issue can be exacerbated in neurobiomedical contexts, where identifying strong and valid IVs can be particularly challenging. Meeting both, the independence and the exclusion restriction assumption can be difficult given the biological complexity and interconnectedness of (neuro)biological systems, such as the brain and the presence of systemic and multiscale brain-body interactions. For example, an IV used to predict GMV might also directly affect HGS. Moreover, the multivariate nature of brain features can make it difficult to identify true paths of influence as the IV could affect different brain regions differently. Consequently, the effectiveness of the IV approach in the case of unmeasured confounders depends on the strength and validity of IVs which are difficult to find for multidimensional, multicollinear and causally intertwined data, such as neurobiological data.

It is important to highlight here once again that while in treatment effect estimation IVs are used to identify the causal effect of a treatment (e.g. drug assignment) on an outcome in a population, the goal in supervised ML deconfounding is to train a predictive model that avoids spurious associations due to confounding - improving generalizability and biological meaningfulness of models. Thus, while the statistical machinery is similar, the objectives diverge: unbiased function approximation in ML vs. estimating e.g. average treatment effects in causal inference.

##### 2.2.2.3. Two proxies

Another strategy for debiasing SML models arising from unmeasured deconfounders - such as *sex-hormone levels* in the prediction of HGS from GMV - is the use of proxy variables. A proxy P is a third variable that is causally influenced by a variable Z, here e.g. the unmeasured deconfounder, but does not itself directly affect the target or the feature(s) (**Box 1**). For instance, in the GMV-HGS prediction, potential proxies for *sex-hormone levels* include *voice pitch (F*_*0*_*), waist-to-hip-ratio (WHR), facial hair* or *ratio of index finger to ring finger length (2D:4D)* - all of which are known to be biologically modulated by testosterone levels (**Figure 3**, orange). Miao et al. ^32^ formalize the conditions under which it is possible to use at least two proxies P1 and P2 to nonparametrically recover the influence of the unmeasured deconfounder Z. This method requires three key assumptions:

###### 1) Conditional Independence

The proxies must be statistically independent of each other when the latent deconfounder Z is held constant.

- Formally: P(P1, P2 ∣ Z) = P(P1 ∣ Z)⋅P(P2 ∣ Z)
- Intuition: Any statistical association between, for example, *voice pitch* and *facial hair* should be explainable solely by shared dependence on *sex-hormone levels*. This assumption is biologically plausible since these traits arise from distinct physiological pathways—laryngeal development vs. follicular activation – with no evidence of a direct causal link between the two.

###### 2) Relevance Condition

Each proxy must provide nontrivial information about the unobserved deconfounder Z.

- Formally: P(P∣Z=z_1_)≠P(P∣Z=z_2_) for some z_1_≠z_2_ (The conditional distribution P(P∣Z) must change as Z changes)
- Intuition: Changes in Z must induce systematic and detectable variation in the probability distribution of each proxy. For example, testosterone levels during puberty lower voice pitch by growth and thickening of vocal folds (Harries et al., 1998) and stimulate facial hair growth via androgen receptor activation (Randall, 2008), i.e. both proxies respond systematically to changes in the deconfounder. In contrast, a variable like *2D:4D*, while causally downstream, it is only weakly associated with prenatal testosterone exposure ^33^. This weak and noisy variation may thus make it fail to satisfy the relevance condition.

###### 3) Rank Condition (generalization of earlier frameworks ^34^ that required functional links between proxies and Z)

The joint distribution of the proxies varies sufficiently across values of Z to allow for its reconstruction up to a transformation.

- Formally: The covariance matrix (conditional expectation operator) of the proxies given Z must be of full column rank. This guarantees that the mapping Z→(P1, P2) is injective, so that different values of Z produce distinguishable configurations of proxy values.
- Intuition: Both proxies are independent noisy reflections of Z, so that their combination uniquely pins down the value of Z. For example, *voice pitch* and *facial hair* reflect testosterone activity via partially distinct pathways, i.e. anatomical and functional independence of the larynx and facial hair follicles. This suggests that their joint variation provides non-redundant information about *sex-hormone levels*, making the rank condition plausibly satisfied.

A central challenge of proxy-based deconfounding is that the three conditions cannot be empirically verified in a data-driven way, since the deconfounder Z is unobserved. For example, conditional independence tests between *voice pitch* and *facial hair* (e.g., partial correlations, conditional mutual information) require conditioning on the unmeasured *sex-hormone levels*. Using a cause of the unobserved deconfounder as a surrogate variable (e.g. *hormone intake*) (**Figure 3**) might appear as a good strategy for data-driven testing of proxy quality, but is conceptually flawed. For example, *hormone intake* is a parent of *hormone levels*, not a noisy measure thereof, and thus violates the structure required of proxies. Even if *hormone intake* were a proxy, it would not be valid to empirically assess the conditional independence of two proxies based on another proxy. Similarly, the relevance and rank conditions depend on the structure of the mapping from Z→P, which requires direct observation of Z. Consequently, the credibility of proxy-based deconfounding must rest on domain knowledge and theoretical justification. This includes extending the DAG from step 1 to incorporate candidate proxies by justifying each edge and the plausibility of the three conditions based on known biology. For instance, one can argue that *voice pitch* and *facial hair* fulfill all three conditions to serve as proxies for *sex-hormone level* given established physiological mechanisms, whereas *2D:4D* may fail the relevance criterion due to weak variation.

##### 2.2.2.4. Broader reflections on handling unmeasured confounders

Despite their appeal, the success of strategies such as front door-adjustment, IVs and proxies hinges on the availability of high-quality variables. For example the IV approach requires strong instruments V to avoid high variance and imprecise estimates. Proxy approaches in contrast risk bias if the proxies fail to adequately capture the unmeasured deconfounder’s information.

Beyond variable quality, the validity of these strategies depends entirely on correct specification of the underlying causal structure. Mischaracterizing causal roles or dependencies can lead to biased estimates, undermining the very purpose of deconfounding. This emphasizes a central point: no deconfounding strategy can rescue missspecifications of causal relationships. If the DAG fails to reflect the data-generating process, all derived adjustments risk introducing rather than mitigating bias.

As a result, in practice, it is often advisable to favor the simplest deconfounding approach, i.e. where the least assumptions must be fulfilled, making the backdoor criterion an attractive choice, if the respective variables are available. The list of discussed alternative strategies is not exhaustive but reflects approaches that are (i) grounded in causal theory, (ii) transparent in their assumptions, and (iii) reasonably implementable in real-world datasets.

### 2.3. Step 3 – Statistical evaluation and adjustment

#### 2.3.1. Statistical evaluation

In the GMV-HGS prediction example, the unmeasured deconfounder *sex-hormone levels* can be circumvented by using an alternative set of backdoor-justified deconfounders, namely *muscle-mass* and *sex* instead of *age* and *sex-hormone levels* (**Figure 3**, orange). Both alternatives are availably measured in the UKB at the same time point as HGS and GMV. and causally relevant as per the DAG (step 1).

In addition to causal relevance, deconfounders need to be statistically associated with both feature(s) and the target as causal relationships only become actionable when they manifest in the data ^35^. We nonetheless put the causal analysis first because this allows for a bottom-up identification of deconfounders (as classically pursuit in statistics) in contrast to a top-down pre-definition of confounders which can create insecurities about what variables to include as confounders (see step 1).

The threshold for what qualifies as sufficient statistical association depends on context, comparable to no hard threshold in for example nul hypothesis testing (convention of p<.05 or p<.01). In our example, *sex* correlates^2^ with HGS at r=.75 and with GMV at r_median_=-.12; *muscle mass* correlates with HGS at r=.76 and with GMV at r_median_=-.08 (**Figure 1c**). The relatively low median correlations between the deconfounders and GMV arise from the multi-dimensionality of the GMV features (1088 brain regions), many of which show heterogenous associations – positive in some regions, negative in others - resulting in a median near zero. This is important to note as statistically unrelated deconfounders should not be adjusted for. In the best case such adjustment would be irrelevant but in the worst case it can introduce bias by leaking information from the deconfounder into the feature or target in the adjustment process ^36^. Lastly, there also must be a statistical association between GMV (X) and HGS (Y) (**Figure 1c**) to assure predictability (even though potentially biased) and thereby debiasing (deconfounding) meaningful.

#### 2.3.2. Deconfounding using linear residualization

Once deconfounders are identified through both causal justification and empirical association (in our case: *sex* and *muscle mass*), models can be adjusted. In SML several (post-hoc) confounder mitigation strategies exist. One of the most established approaches is linear residualization, where confounder information is mass-univariately linearly regressed out of features or the target (e.g. ^37^) (residualization). In our example, we residualized the GMV features for the identified deconfounders *muscle mass* (operationalised as lean mass) and *sex*. Using a linear SVR (L2) model trained on residualized features, we observed no correlation between true and predicted HGS (r=0.00, R^2^<0^3^; **Figure 1b**, right) (for methods see supplementary materials).

This constrasts strongly with the unadjusted model, which yielded r=.48 (R^2^=.09) between true and predicted HGS (section 2). Keeping in mind the combination of a simple linear SVR with linear confounder regression, the collapse in predictive performance after deconfounding implies that the earlier performance was largely driven by confounding bias. In other words, the unadjusted model did not learn meaningful biological relationships between GMV and HGS but rather exploited demographic and behavioural correlates. Such a model would likely fail in datasets with different distributions of *sex* or *muscle mass*, undermining both generalizability and scientific insights.

On the other hand side, the poor performance of the unbiased model does not necessarily imply the absence of any meaningful GMV-HGS relationship. Instead, it signals that a simple linear model may be insufficient to capture more nuanced, biologically plausible patterns in the data. Further exploration using non-linear models or multimodal inputs may be required to recover valid signal under proper deconfounding constraints.

## 3. Limitations of linear (feature) residualization and alternative approaches

### 3.1. Limitations and strength of linear feature residualization

Linear residualization of features is commonly used in neurobiomedical predictive modelling for confounder adjustment. Despite its popularity, it has two main limitations relevant to this work, namely (1) the assumption of a parametric linear relationship between confounders and each feature and/or the target, and (2) the adjustment typically being applied to features or target, not both.

First, linear residualization effectively only removes linear confounding effects. While this may be sufficient with linear predictive models, it becomes problematic with non-linear prediction algorithms. These can leverage residual non-linear confounding information, resulting in biased predictions despite linear adjustment.

Second, standard practice in SML often involves residualizing either the features or the target, but rarely both. Typically, only features are adjusted to preserve interpretability of the target, which often represents the neurobiological entity of interest (e.g. diseases status, cognitive score). In the following, we elaborate why one-sided confounder adjustment can be of concern.

#### 3.1.1. On the potential benefits of feature and target adjustment – signal contribution perspective

Linear residualization of only the features X implicitely assumes that all variation in the target Y that is associated with confounders Z is fully captured by X. In practice, this assumption rarely holds. While residualizing X removes the confounders’ variance from X (“clean” features), it leaves confounder-related variance in Y. Thus, Y is still partially influenced by Z so that the statistical relationship between X and Y becomes misaligned: the model attempts to predict Y from features that no longer carry the Z-related information, while Y still contains it. This misalignment can have two main consequences. First, the one-sided adjusted model may remain partially biased. Since confounding variance remains in Y, the model may over- or under-estimate relationships between X and Y, depending on the structure of Z’s effect. Second, predictive accuracy may be reduced because the remaining information in residualized X cannot explain the signal in Y linked to Z. In effect, while there is no formal guarantee that residualizing both X and Y is benefitial, it could improve statistical alignment between features and target, supporting potentially less biased and more accurate predictions.

#### 3.1.2. Necessity of feature and target adjustment - causal perspective

As confounding is inherently a causal concept, causality-based reasoning offers a deeper lens to understand the limitations of only feature or only target residualization. Previously, we introduced the backdoor criterion as a method solely for confounder identification based on a DAG. But it does more than this: Being rooted in causal literature, it provides a formal basis to estimate the interventional causal effect of X on Y – expressed as P(Y∣do(X)) (Box 2).

This interventional distribution represents the probability of observing Y=y when actively intervening to set X=x in the population. In contrast, the potentially confounded observational distribution P(Y|X) reflects the probability of observing Y=y among individuals for whom X=x is observed, regardless of other influencing factors. For instance, P(HGS=29kg|GMV_anterior_globus_pallidus_=300mm^3^) ^4^ might conflate effects of sex or body composition, whereas P(HGS=29kg|do(GMV_anterior_globus_pallidus_=300mm^3^)) isolates the interventional causal influence of GMV (**Box 2**).

According to the backdoor adjustment formula ^38^, for variables Z that satisfy the backdoor criterion, the interventional distribution can be computed as:

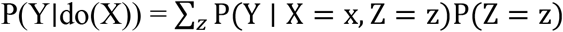

Here:

- P(Y∣X, Z=z) is the conditional observational distribution, i.e. how likely it is to observe a certain value Y, given that a certain value X=x and Z=z was observed (e.g., P(HGS=29kg|GMV_anterior_globus_pallidus_=300mm^3^, sex=female)), and
- P(Z=z) is the marginal probability distribution of Z (e.g., P(sex=female) = 0.6 if 60% of the population are female)

This formula tells us that to estimate the interventional causal effect of X on Y, we must assess the conditional probabilities P(Y∣X=x, Z=z) across all levels of Z (e.g. male, female), then aggregate them using the prevalence of each z in the population as weights. For example, the causal effect of a GMV of 300mm^3^ in the left anterior globus pallidus on HGS would be obtained by summation (discrete Z) or integration (continuous Z) of the weighted conditional distributions over all sexes: P(Y ∣ do(X = 300)) = ∑_*z*∈{*male,female*}_ P(Y ∣ X = 300, Z = z)P(Z = z).

Evaluating each level of the confounding variable Z separately is essential. By holding Z constant, its influence on the X-Y relationship is effectively neutralized. This ensures that any observed variation in Y can be solely attributed to changes in X, comparable to the strategy of matching samples. Moreover, by aggregating across all levels of Z using a weighted sum (or integral), the approach estimates the effect of setting X=x in the entire population, rather than being limited to the subpopulation for which X=x is naturally observed. Together, by conditioning on Z, this strategy eliminates variation in the X-Y relationship from shared dependence on Z, effectively blocking information flow along the non-causal backdoor path from X to Y via X ← Z →Y. As a result, any remaining association reflects the direct interventional causal influence of X on Y.

In predictive SML, we approximate this idea using residualization. Instead of adjusting additively as in the backdoor formula, residualization removes the influence of *Z* subtractively by regressing it out from both X and Y. This yields:

- 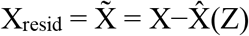 (features orthogonal to (independent of) Z)
- 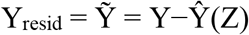 (target orthogonal to (independent of) Z),

with 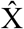 and Ŷ being the respectively predicted values of X and Y from Z. A model is then trained to predict 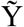 from 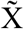. This procedure removes Z-related variation from both the features and the target, i.e. cutting off both Z → X → Y and Z → Y, so that the remaining signal in the model now reflects the component of X that explains variation in Y independent of the confounders Z. In this sense, dual residualization (adjusting both features and target) enables not just proper model debiasing, but could additionally provide the opportunity to examine causal effects of X on Y using predictive models. However, as discussed below, the conditions under which causal claims from deconfounded SML models are valid must be carefully examined.

#### 3.1.3. Reasons linear (feature) residualization is used despite limitations

Despite well-founded arguments for residualizing both features and the target and for using non-linear models for confounder adjustment, linear feature residualization remains common, especially in fields such as brain-behaviour predictive modelling. One reason is its simplicity: assuming linear confounder-feature/target relationships limits overfitting risk, avoids hyperparameter optimisation, and makes the residualization easily integratable in SML cross validation (CV) pipelines without leakage. This makes it a practical choice for achieving debiased predictions.

Another key reason is that residualizing only features preserves interpretability of the target, which is typically the scientific or clinical interest (e.g. disease status, cognitive score, age). Residualizing the target can obscure its meaning. In such cases, quantifying confounder-target associations (see section 2, step 3) can support informed decision and transparent communication of the trade-off between confounding influence and target interpretability.

Additional reasons include established practice, limited tooling, and the sometimes implicit misconception that cofounders affect the target only via features (Z→X→Y), ignoring direct Z→Y pathways. More broadly, while linear feature residualization is an established convention, a causal approach to confounding is less widespread in certain SML communities. Finally, clear, accepted alternatives are scarce. While alternatives such as double/debiased machine learning (DML) ^39^ can offer more thorough bias removal, they are designed for causal inference and are not (yet) adopted for debiasing prediction models.

### 3.2. Double/Debiased machine learning to overcome the limitations of linear feature residualization for debiasing SML models?

Double/Debiased Machine Learning (DML) ^39^ is a method to estimate causal parameters, such as average treatment effects in the presence of high-dimensional confounding. It is both theoretically well established and practically implemented in various Python and R based toolboxes such as EconML (e.g. ^40^) or DoubleML (e.g. ^41^). While we leverage causal tools to unbias SML models, they leverage SML tools (correlative) to obtain unbiased causal parameter estimates. Even though originally developed for causal questions, DML provides valuable insights that may be repurposed to improve confounder adjustment in supervised machine learning (SML) under theoretical adaptation and practical considerations.

#### 3.2.1. DML – the method

The DML framework targets the estimation of a causal parameter *θ*_0_ from data W = (Y, D, X_C_), where Y is an outcome, D a treatment variable and X_C_ a potentially high-dimensional set of confounders^5^. The relationship between variables is described by two partially linear models:

1. Treatment/Confounder-Outcome relationship:

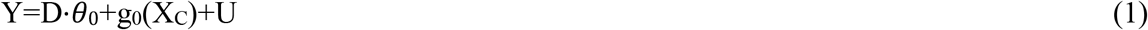

with stochastic errors U, following E[U∣D,X_C_]=0
2. Treatment-Confounder relationship:

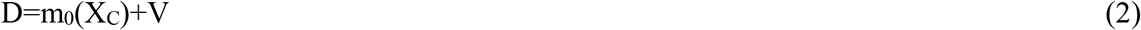

with stochastic error V, following E[V∣X_C_]=0

While the treatment-outcome relationship is assumed to be linearly described by the causal parameter *θ*_0_, the nuisance functions g_0_ and m_0_ that model the respective relationship with the confounders can be high-dimensional, non-linear and complex, hence the suggested usage of regularised SML tools, such as LASSO or *l*_+_ -penalized neural networks, to get an estimate of g_0_, i.e. ĝ_0_. Regularised algorithms are useful to resolve bias-variance trade-offs in a prediction context but would lead to a biased estimate of the regression coefficient *θ*_0_ if the prediction ĝ_0_ would be directly plugged into (1). To counter this, DML introduces a cross-fitting strategy, splitting the data into an auxiliary and main subset: The nuisance functions m_0_ and g_0_ are learned on the auxiliary data and treatment D and outcome Y are residualized 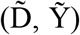 with their predicted values 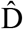 and Ŷ obtained using (2) and (1), respectively. 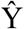 and 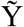 are orthogonal (independent) to X_C_ and can therefore be used to get a debiased estimate *θ*_0_ on the separate main data, ensuring so-called Neyman orthogonality. This orthogonality condition is key to robustness, as it guarantees that small errors in nuisance estimation ĝ_0_ and 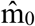 do not substantially bias the estimate *θ*_0_.

#### 3.2.2. Transferring the DML approach to debiasing SML models – challenges and opportunities

Directly translating DML framework into the SML context poses nontrivial methodological and practical challenges^6^. Fundamentally, the objectives differ: while DML aims to recover a treatment effect *θ*_0_, SML seeks to build predictive models of Y that are robust to confounding.

Methodologically, an essential aspect of DML’s theoretical foundation lies in its use of sample splitting into auxiliary and main datasets (or generalized to k-fold), which allows the scaled estimation error of the target parameter *θ*_0_ (i.e 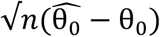) to be decomposed into three terms which under certain constraints vanish asymptotically. Specifically, for the third term in this decomposition (equation 1.6 in Chernozhukov et al (2018):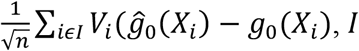 : main sample split of size *n*) its probability vanishes due to the use of the sample-splitting strategy. However, this result has only been rigorously proven for the partially linear model. In contrast, SML typically seeks to estimate complex, potentially non-linear relationships between the target Y and the features X. Therefore, before DML can be validly applied in SML contexts, it would be necessary to establish that the required assumptions still hold under such general, non-linear settings.

While the features X in SML are seen as the conceptual anologous of the treatment D in DML, dimensionality roles are reversed. DML is developed under the assumption of a binary treatment D and high-dimensional confounders X_C_, whereas in SML the roles are reversed: features X are high-dimensional, and confounders C are few, which can fundamentally alter the estimation landscape. Although the original DML framework allows some flexibility in dimensionality of the treatment variable (footnote 1 in Chernozhukov et al (2018)), using a high-dimensional feature matrix X as a “treatment” analogue can introduce complications. With high-dimensional features X, such as 1088 brain regions in the GMV-HGS example, modelling the confounder-feature relationship involves predicting multivariate targets because the features of the actual prediction aim become the targets of the confounder-feature modelling^7^. This presents two modelling options: (1) fitting a multi-output model with e.g. 1088 targets and only few inputs (e.g. the confounders *sex* and *muscle mass*), or (2) training e.g. 1088 separate models in a mass-univariate fashion. While the latter is common in typical linear feature residualization, more complex models require different ML strategies such as nested cross-validation and are more prone to overfitting. Crucially, if the orthogonalization step that relies on residualizing X and Y is not implemented with sufficient precision due to model misspecification or overfitting, then the cross-fitted bias term (equation 1.6. in Chernozhukov et al (2018)) might not vanish as expected, leaving residual bias even after cross-fitting.

Adopting DML to the SML context practically introduces challenges to model training. The DML procedure only requires cross fitting for confounder modelling but uses a linear regression for treatment effect estimation. In contrast, SML relies on complex models for predicting Y from X that already require nested cross-validation. When now feature and target residualization additionally require complex ML models which each need nested cross-validation, the resulting data partitioning becomes intricate. Concretely, it requires a first split into auxiliary and main folds (as per the DML framework), and then internally within the auxiliary and main fold further splits for nested cross-validation for hyper parameter optimisation (inner loop) and estimation of generalization error (outer loop) of all nuisance models (in the auxiliary fold) as well the actual SML model (in the main fold) (**Figure 4**). This hierarchical structure not only sharply reduces the effective sample size available for training and adds substantial computational burden, it also increases susceptibility to data leakage, especially when additional preprocessing or feature selection steps are needed.

**Figure 4.**
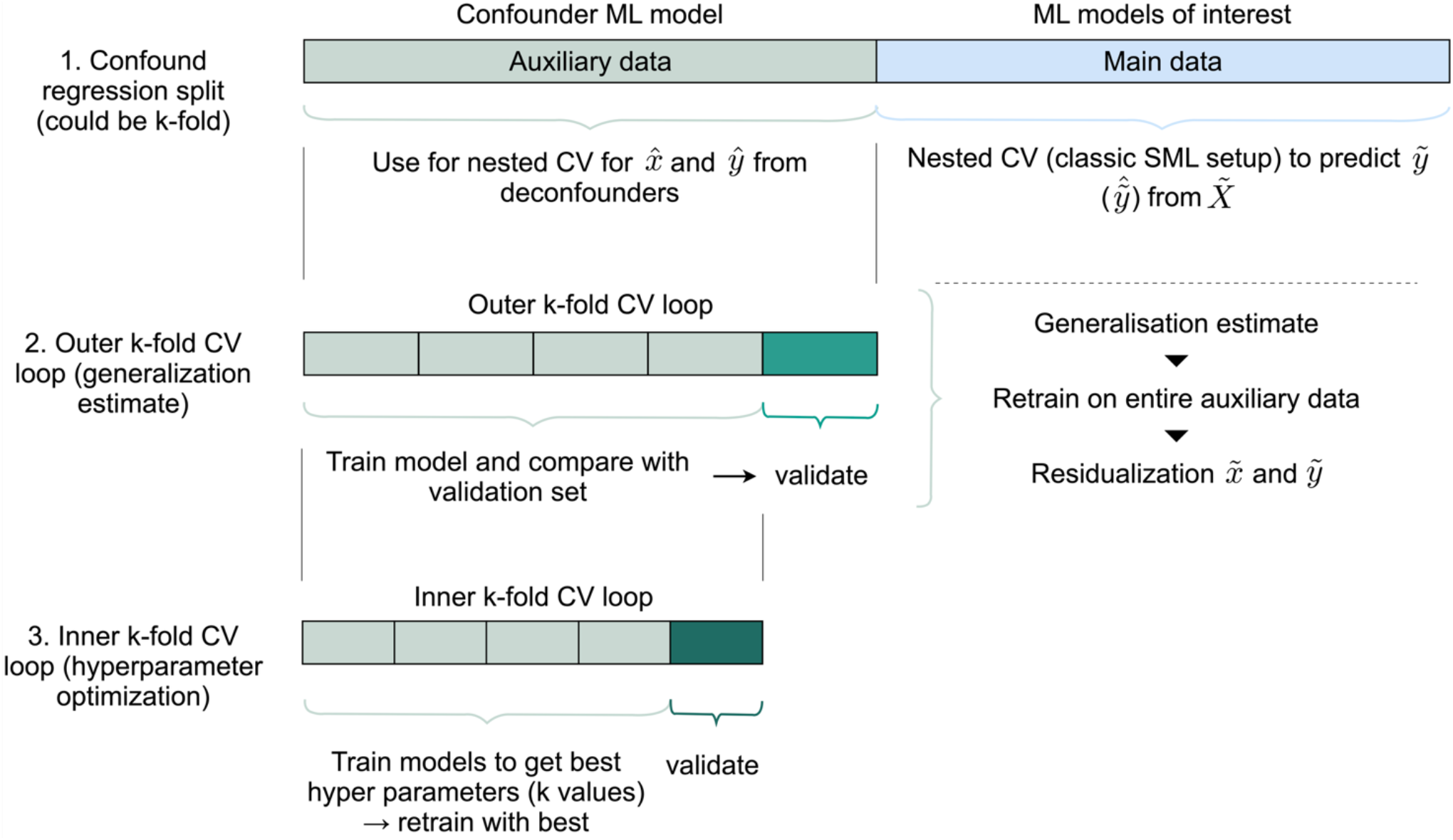
Theoretical cross validation scheme in case of the application of core DML principles to SML. Illustration of the hierarchical data partitioning required when combining Double/Debiased Machine Learning (DML) sample splitting with nested cross-validation (CV) for supervised machine learning (SML). First, the dataset is split into auxiliary and main folds according to the DML framework, enabling cross-fitting of nuisance models on the auxiliary data to prevent overfitting. Within each fold, further nested CV is required: an inner loop for hyperparameter optimization and an outer loop for out-of-sample performance estimation of both the nuisance models (in the auxiliary fold) and the main SML model (in the main fold). This multi-level split could support proper residualization of features (and targets) by enabling more complex models for confounder adjustment through limitating the risk of overfitting. However, it substantially reduces effective sample size, increases computational cost, especially when the auxiliary-main split is also extended to k-fold splits. Additionally, it raises the risk of data leakage if additional preprocessing or feature selection is performed.

Finally, the conceptual concerns remain regarding target residualization and interpretability. In causal inference, residualizing the outcome is unproblematic because the causal estimand *θ*_0_ is the parameter of interest. In SML, however, the original target is the quantity of interest, such as HGS in the above example, not some derivative thereof. Hence, it is problematic that residualized targets are not directly neurobiologically interpretable.

Despite these challenges, adapting DML principles to the SML setting opens promising avenues for improving confounder-adjusted predictions. First, it strengthens the rationale for residualizing both features and the target – a deliberate trade-off between complete removal of confounding information and preservation of target interpretability. Second, DML provides a mathematically well-theorised framework for employing flexible ML estimators in residualization, extending beyond simple linear regression. Third, the cross-fitting scheme central to DML highlights the need for separate data partitions to prevent overfitting of complex nuisance estimators. SML pipelines do rely on data-splitting through nested cross-validation for hyperparameter optimisation (inner loop) and out-of-sample performance generalization estimation (outer loop). However, confounder adjustment is conducted within each fold of the innermost data split rather than on independent partitions. Incorporating an additional cross-fitting step (**Figure 4**), inspired by DML, could therefore enhance SML practice by yielding out-of-sample confounder-adjusted feature residuals, ensuring robustness of confounder adjustment models to overfitting even when complex models are employed.

In summary, the DML framework provides a powerful tool for debiased causal estimation, with potential to inspire more thorough confounder adjustment in SML. However, its direct application to building debiased SML models requires further theoretical and practical adaptations.

## 4. Positioning of deconfounded SML between predictive modelling and causal inference

Deconfounding supervised machine learning (SML) is necessary to mitigate bias in predictive models but additionally can bring SML closer to causal inference. SML is fundamentally associative, capturing statistical patterns based on conditional, marginal, or joint distributions. However, sufficient deconfounding, e.g. by drawing inspiration from double machine learning (DML), could enable causal interpretation in SML models. Therefore, the question arises: how far can SML go beyond prediction toward interpretable causal claims?

To contextualize the question, a recap of basic causal inference concepts is helpful. Accordings to Pearl’s ladder of causation, SML operates at the lowest, associational rung (e.g. P(Y|X)), whereas causal inference aims for higher rungs, namely interventions (P(Y|do(X))) or counterfactuals, which require additional structural assumptions ^26,42^. Central to causal inference is treatment effect estimation, either at the individual level (ITE) or averaged across a population (ATE). Randomized control trials (RCTs) enable causal identification via randomization, ensuring exchangeability/ignorability (no unmeasured confounders). When RCTs are unfeasible frameworks such as Structural Causal Modeling (SCM) ^43^, Rubin’s potential outcomes framework ^44,45^ and Structural Equation Modeling (SEM; ^46^ offer alternatives. SCM, in particular, fromalizes causal assumptions about the data-generating processes via DAGs, enabling identification of confounders, mediators, and colliders.

### 4.1. Distinction of causal Machine Learning from debiased SML

Causal machine learning (causal ML) aims to bridge ML and causal inference, shifting focus from prediction to estimating interventional effects. For example, instead of predicting diabetes risk, causal ML might estimate how a lifestyle intervention changes diabetes risk.

As outlined by Feuerriegel et al. ^47^, causal ML typically involves:

1. Problem setup: Defining the causal question quantity of interest (e.g., ITE/ATE, Conditional ATE) and relevant variables (treatment, confounders, outcome) and assessing the plausibility of assumptions;
2. Choosing and fitting the causal ML methods (e.g. causal forest, meta-learners, Bayesian networks).

Even though the problem setup evaluates plausibility of assumptions, integrating an explicit step to model causal variable relations would further strengthen the selection of relevant variables and ensure appropriate confounder adjustment (here: chapter 2). Beyond causal ML for treatment effect estimation, newer directions include counterfactual data generation e.g. through deep learning and causal graph discovery to learn DAGs from data to complement expert-driven causal modelling.

Despite these causal ML applications, the question remains, if and how debiased SML could allow for causal insights not in the sense of estimating treatment effects but in the sense of understanding causal feature-target mechanisms. For instance, rather than associative prediction of HGS from parcellated GMV, causal SML would ask if the GMV in the left anterior globus pallidus is a causal driver of HGS, aiming for biological mechanisms rather than statistical associations.

### 4.2. Assumptions required for causal interpretability and positioning of debiased SML

To evaluate the potential of SML for causal insights, key assumptions from causal inference must be considered and evaluated whether they can be met in debiased SML models, particularly in neurobiological contexts.

#### 1. Ignorability

All confounders must be observed and accounted for. Our DAG-guided framework and DML discussion aims for this, though high-dimensional, intertwined neurobiomedical data and methodologcial limitations make full coverage challenging.

#### 2. Causal Markov Assumption

Variable must be conditionally independent of their non-descendants given their direct causes. For instance, HGS is conditionally independent of *sex hormones* given *muscle mass* (**Figure 3**). This assumption must be assumed in expert-defined DAGs, but biological systems may require or inadvertently contain (hidden) cycles, undermining causal interpretation.

#### 3. Positivity

All levels of the treatment (features, e.g. low/high GMV) must occur across all confounder strata (e.g. young/old). For instance, usually low GMV is particularly associated with older age, but not observed in young adults, so it cannot be identified what would happen if a young person had low GMV, violating positivity. In ML terms, this corresponds to poor extrapolation, covariate shift or domain mismatch challenges. Even a deconfounded model cannot learn a causal relationship in regions of the feature space that are unobserved or underrepresented.

#### 4. Faithfulness (Absence of Coincidental Independencies)

All observed conditional independencies (**Box 2**) in the data correspond to those implied by the structure of the causal DAG and are not the result of numerical coincidences or cancellation effects. Faithfulness links statistical patterns to causal structure. Faithfulness would for example be violated in a setup with GMV-> HGS, physical activity-> HGS and physical activity-> GMV, if the effects of GMV-> HGS and physical activity-> HGS are equal in magnitude but opposite in direction. The net observed association between GMV and HGS could be close to zero, despite a true underlying causal pathway. This creates a conditional independence in the data that does not match the actual DAG. Sometimes unfaithfulness can be prevented by confounder correction, but generally is a bigger concept, ruling out all statistical independencies that result from exact numerical cancellations, not just dues to confounidng. Although strong and ultimately untestable, faithfulness is often regarded as a reasonable default assumption—unless there is domain-specific reason to suspect causal cancellations.

#### 5. Consistency

An individual’s observed outcome must equal their potential outcome under a certain treatment. In our context, if a person has a specific GMV level, their observed HGS is assumed to reflect what would occur under that same GMV level in a counterfactual scenario. Additionally, it assumes no interference between individuals, akin to the independent and identically distributed (i.i.d.) assumption in ML. Violations occur when identical treatment/feature values (e.g., GMV) arise from different biological processes – such as lifelong physical training in one person versus genetic predisposition in another – potentially leading to different outcomes (HGS) despite the same observed GMV. This illustrates a key limitation of applying debiased SML for causal insight in complex biological systems with latent heterogeneity.

### 4.3. How much causal claims do properly debiased SML models allow for?

Given the discussed constraints, how far can a well-deconfounded SML model support causal interpretation of the relationship between X and Y? Consider a well debiased SML model: Relevant confounders are correctly identified, features and the target are appropriately residualized (linearly and non-linearly), and predictive performance is high. Can we then conclude that X causes Y?

Not necessarily. While such a model may hint towards a causal relationship, it does not guarantee one. Violations of assumptions, such as unobserved confounding (violating ignorability), unaccounted DAG cycles (violating the causal Markov condition), or features reflecting proxies for latent biological processes (violating consistency), can still hinder a causal interpretation.

Even if all assumptions hold, interpreting the learned association P(Y∣X) as the interventional distribution P(Y∣do(X)) remains problematic due to ambiguity of causal direction. SML models estimate P(Y∣X), which is direction-agnostic^8^. For instance, predicting blood pressure (Y) from drug dosage (X), reflects learning in causal direction (X → Y), while predicting disease status (Y) from brain structure (X) may reflect learning in anti-causal direction (Y → X) - yet in both cases, the model is trained to predict Y from X ^48^.

In the GMV-HGS example, predictions by a SML model may reflect GMV affecting HGS (e.g. motor cortex GMV determines strength) or HGS influencing GMV (e.g. strength training induced neuroplasticity). Both pathways are biologically plausible. Moreover, high-dimensional models, such as the exemplary one using 1088 GMV parcels, likely capture mixtures of causal, anti-causal, and non-causal patterns, reflecting the dynamic, bidirectional nature of brain-behaviour relationships. Disentangling these directions requires additional information: domain knowledge, experimental interventions, or further modelling assumptions such as additive noise models, invariance, or algorithmic asymmetries (see e.g. work from Schölkopf et al. for details).

In summary, debiased SML models can offer insights consistent with causality, but not proof of it. In addition to proper deconfounding, causal assumptions must be satisfied and the model’s predictive direction must align with the true data-generating process. Because SML inherently learns P(Y∣X), interpreting this as P(Y∣do(X)) requires a strong justification without which any causal interpretation remains suggestive rather than definitive.

## 5. Conclusion

Confounding is a major source of bias in neurobiomedical supervised predictive modeling, and principled confunder selection - beyond correlation- or heuristic-based justification - is essential for meaningful predictions. We propose a three-step framework for confounder selection - causal analysis via DAGs, guidance for appropriate deconfounder selection strategy, and statistical validation - that integrates tools from causal inference into associative supervised machine learning (SML). While linear (feature) residualization is commonly applied for confounder adjustment, it is limited in handling complex, non-linear confounding and may leave residual confounding signal. Approaches such as Double/Debiased Machine Learning can inspire improved adjustments in SML workflows but theoretical and practical challenges remain. Importantly, even with appropriate deconfounding, SML remains fundamentally associative, and causal interpretations would require additional assumptions. Nonetheless, properly deconfounded models are critical for generalizable and neurobiomedically informative supervised predictive models, forming an indispensable foundation for neurobiomedical ML research.

### Box 1 Types of third variables and associated biases

When investigating the relationship between a predictor (feature) X and an outcome (target) Y, third variables can be related to X and Y in different ways. The different natures can be best visualized by using directed acyclic graphs (DAGs) (**Box 2**).

**Types of third variables**

A **confounder Z** is a (direct or indirect) common cause of the feature X and the target Y (Fig. B1.1A). Formally, a confounder can be defined as a variable Z that leads to a discrepancy between the conditional probability of Y given X (*seeing*) and the probability when intervening on X (*doing*): *P*(*Y* | *X*) ≠ *P*(*Y* | *do*(*X*)). Not controlling for a confounder will obscure the causal effect of X on Y. One can either control for the confounder itself or any variable that lies on the path X ← Z → Y.

A **collider C** is the common effect of a feature X and a target Y. Conditioning on a collider induces a spurious (i.e. non-causal) association between X and Y (Berkson’s paradox) ^21,23,49^ (Fig. B1.1B). In other words, if X and Y were independent to begin with, conditioning on Z will make them dependent. (see Berkson’s paradox for an example).

A **mediator M** is caused by X and is a cause of Y ^50–52^ (Fig. B1.1C). For example blood pressure might mediate the relationship between a drug and the risk for a heart attack such that the drug decreases the risk for a heart attack via lowering blood pressure. When interested in the total effect of the predictor on the outcome (X → Y and X → Z → Y), conditioning on M blocks the causal path X → M → Y and will hence only reveal a partial effect. When only interested in the direct effect X → Y conditioning on M can nonetheless lead to biased estimates, if the mediator and the outcome share a common cause because then the mediator is a collider for the predictor and this common cause.

**Fig. B1.1.**
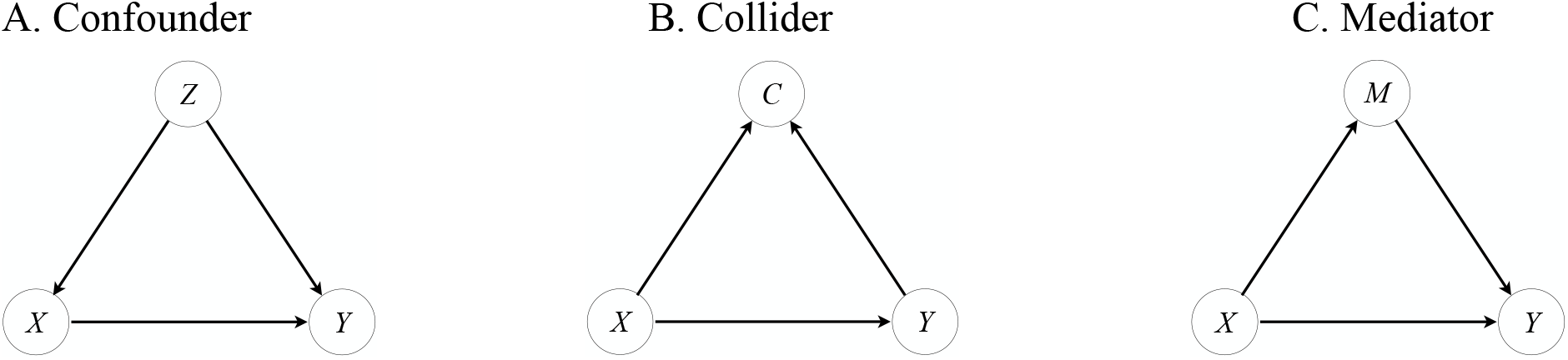
DAG of a confounder (A), a collider (B) and a mediator (C).

*Note*: Defining confounding via correlations and not as a causal note is not sufficient because each of the causal structures A.-C. produces a correlation between the third variable and both X and Y, which could all produce the same correlation matrix. Consequently, correlations cannot help to distinguish between a confounder, a collider or a mediator ^24,25^.

A **proxy P** is caused by X but has no causal relation to Y ^38^ (Fig. B1.2A, B). If the feature X is a perfectly reliable measure of the construct of interest, then controlling for a proxy will not affect the path X → Y. However, in many disciplines X is an unreliable measure of the true causal variable, e.g. a MRI scan for the underlying morphology. In this case, the proxy is a second unreliable measure of the same true predictor (e.g. morphology) and conditioning on this proxy will partition the true predictive effect between the two unreliable proxies so that neither of the unreliable measures will capture the full causal effect ^18^. The same logic applies for proxies of confounders.

An **instrumental variable I** for the confounded relationship between X and Y needs to fulfill the following criteria (Fig. B1.2.C):

a. Z and I are independent, i.e. there is no arrow between Z and I.
b. There is an arrow between I and X.
c. There is no direct arrow between I and Y, i.e. no direct causal connection.

There are no confounders of the relation between I and Y, so that any observed association must be causal. Likewise, since the effect of I on Y goes through X, one can conclude that the observed association between X and Y must also be causal. An instrumental variable is therefore similar to a coin flip, which simulates a variable with no incoming arrow.

**Fig. B1.2.**
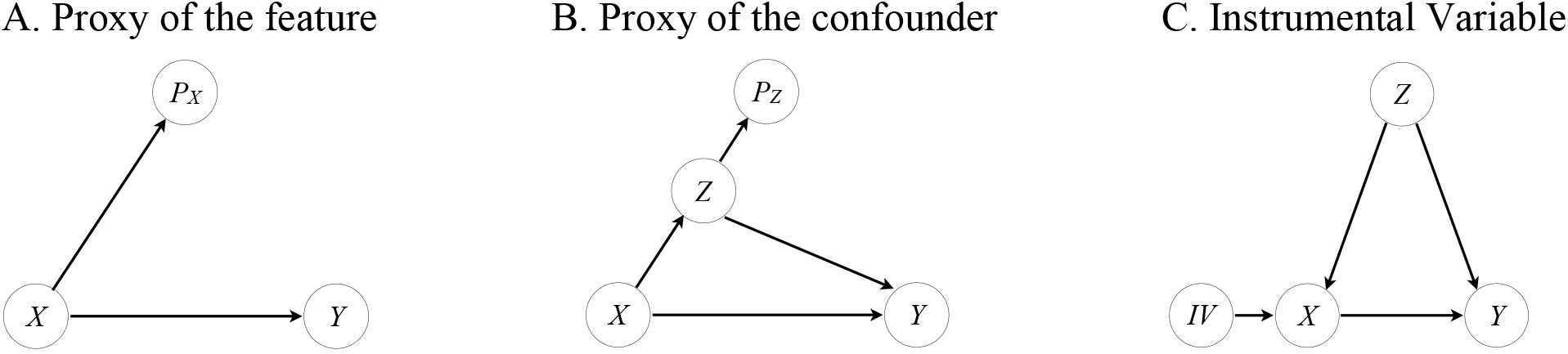
DAG of a proxy of the feature (A) or a confounder (B) and an instrumental variable (C).

**Types of biases (paradoxes) associated with third variables**

Simpson’s paradox (confounder bias)

Simpson’s paradox is a statistical phenomenon in which the statistical relationship between two variables in a population can appear, disappear, or reverse when splitting the population in subgroups or when aggregating two heterogenous subgroups into a population. For example, two variables might be positively associated in the overall population but either not or negatively associated within the subgroups^53^. More generally, it is characterized by the statistical results of the subgroups differing from the aggregated population. It alerts to cases where at least one of the statistical trends (either in the aggregated data, the partitioned data, or both) cannot represent the causal effects^42^.

Berkson’s paradox (collider bias)

Berkson’s paradox is the opposite of the Simpson’s paradox, i.e. it occurs when falsely conditioning on a variable that is the effect of both the feature(s) and the target (collider). Conditioning on such a collider creates a spurious association between the feature(s) and the target. For example, performing a study on patients who are hospitalized, one controls for/conditions on hospitalization. However, if only a disease 1 and a disease 2 together could lead to hospitalization in the first place (with no causal relation between the diseases), conditioning on hospitalization (by performing the study only on hospitalized patients) would introduce non-existing relation between disease 1 and disease 2.

### Box 2 Basic Causal Concepts and Terminology

**Directed Acyclic Graph (DAG)**

Formally, a graph *G* is a collection of nodes and edges that connect (some of) the nodes. In a directed graph the edges are directed, i.e. pointing from one node to another. Visually, arrows indicate the directions. Nodes connected by one edge are called adjacent. A path in a graph is any sequence of adjacent nodes, regardless of direction of the edges that join them, e.g. X ← Z → Y is a path, but not a directed path but X → Z → Y is a directed path. A directed cycle is a directed path that starts from a node X and ends in X. A directed acyclic graph (DAG) is a directed graph with no directed cycles. DAGs obey the local Markov assumption that given its parents in the DAG a node X is independent of all its non-descendants.

Practically, a DAG supports formalization of causal relations or assumptions between variables (nodes), where the arrows (edges) represent directions of known or suspected causal relationships between two variables. For example, X → Y means that X is a direct cause of Y, i.e. the arrow implicitly says that some probability rule or function specifies how Y would change if X were to change. The rule according to which this change happens might either be known (e.g. previous research) or has to be estimated from data. The practicality of DAGs is that they allow to express how a joint distribution over a set of random variables factorizes because they allow to encode (conditional) independence relationships.

**Probabilistic language**

To formally express and intuitively exemplify different types of probabilities we use the neuroimaging example of gray matter volume (GMV) in the left anterior globus pallidus (denoted as G) and hand grip strength (HGS, denoted as H).

1. **Marginal probability**: P(G=g) The probability that a randomly selected individual has GMV G=g in the left anterior globus pallidus. *Example*: What is the probability that a person’s GMV in this region is 400 mm^3^?
2. **Joint probability**: P(G=g, H=h) The probability that an individual simultaneously has a GMV of g and HGS of h. *Example*: What is the probability that someone has a GMV of 400 mm^3^ and a HGS of 35 kg?
3. **Conditional probability**: P(H=h∣G=g) The probability distribution HGS among individuals with a given GMV value. *Example*: Among individuals with a GMV of 400 mm^3^, what is the probability distribution of their HGS? ⥤ associational reasoning based on observation.
4. **Interventional probability (causal)**: P(H=h∣do(G=g)) The probability of HGS if we were to hypothetically intervene and set the GMV to g for all individuals. *Example*: What would the distribution of HGS look like if we could biologically set GMV in this brain region to 400 mm^3^ for everyone? ⥤ causal reasoning under intervention. Corresponds to what is estimated in experimental or simulated interventions.
5. **Conditional independence**: X⊥Y∣Z Two variables X and Y are conditionally independent given a third variable Z if, once Z is known, knowing X provides no further information about Y, and vice versa. *Example*: Suppose physical activity (P) affects both GMV (G) and HGS (H). Then GMV and HGS may appear correlated. However, once physical activity is accounted for, GMV and HGS may become conditionally independent: G⊥H∣P.

### Box 3 Two classic ways to account for confounding influences based on DAGs

There are several ways to identify and account for confounding influences. Two of the most known ones are the backdoor and frontdoor criterion.

<u>Backdoor criterion</u>

A backdoor path is any path from X to Y that starts with an arrow pointing into X, for example X ← Z → Y in Fig. B3.1A. Backdoor paths are *non-causal* paths. To deconfound X and Y one needs to block every *non-causal* path between X and Y without blocking or perturbing causal paths. This can be achieved by adjusting for variables with an incoming arrow into X based on the respective DAG (Fig. B3.1A). These variables are called deconfounders and can differ from the set of confounders, for example when a confounder does not need to be controlled for because the backdoor path is already blocked by a collider (Fig. B3.1B) or when the actual confounder is unmeasured, but the backdoor path can be blocked by controlling for a measured deconfounder (Fig. B3.1C). Randomized control trials (RCTs) avoid confounding as non-causal pathways are blocked through randomization.

**Fig. B3.1.**
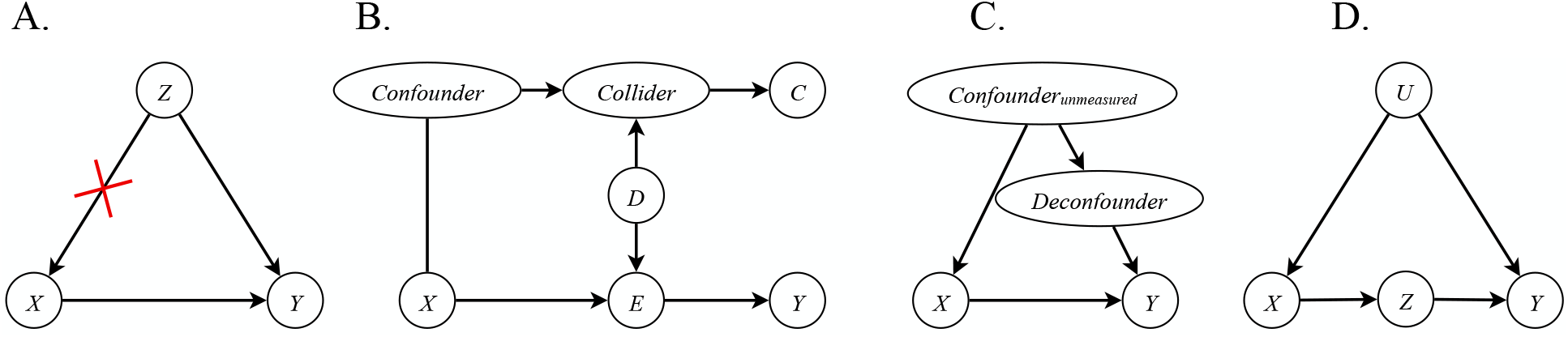
Different positions of a confounder in the DAG structure require different confounder adjustment strategies.

<u>Frontdoor criterion</u>

The backdoor criterion is not feasible when one (or all) deconfounders cannot be measured or are not available. In this case the frontdoor criterion can be applied. The frontdoor criterion (Fig. B3.1D) requires a variable Z that

a) intercepts all direct paths from X → Y
b) there is no backdoor path from X to Z
c) all backdoor paths from Z to Y are blocked by X.

As the relationship between X and Y is confounded by the unobserved variable U, in the frontdoor criterion the effect of X on Y is estimated indirectly by combining the estimate of effect X → Z and of Z → Y.

## Data Availability

This research has been conducted using the UK Biobank Resource under application number 41655.

## 6. Acknowledgments

This research has been conducted using the UK Biobank Resource under application number 41655. This research was funded by the Deutsche Forschungsgemeinschaft (DFG, German Research Foundation) – Project-ID 431549029 - Collaborative Research Centre CRC1451 on motor performance project B05.

## 8. Author Contributions

**VK**: conceptualization, formal analysis, methodology, visualization, writing – original draft, writing review and editing; **CH**: writing – review and editing; **SBE**: funding acquisition, resources; **CR**: writing – review and editing; **FR**: supervision; **KRP**: supervision, writing – review and editing

## Footnotes

1 As we leverage tools from causal inference, we will use terminology such as “X effects/causes Y” and notation such as X→Y, but we do not directly imply that the deconfounded SML model allows for causal claims. We will discuss later in the paper where debiased SML can be positioned w.r.t. causal inference.

2 Point-biserial correlation (see methods in supplementary materials for details).

3 R^2^ refers to the coefficient of determination as commonly used in the ML literature and not to squared correlation values.

4 The probability of observing a HGS (Y) of 29kg (y) in people where the GMV of the left anterior globus pallidus (X) is 300mm^3^ (x).

5 We here stick with the variable naming as used by Chernozhukov et al. (2018), where they use X for confounders and not features like in the SML context. For distinction we label it here X_C_. D would resemble a binary feature (0 or 1) in the SML context.

6 In the following we will adopt the following notation: Y: outcome or target, Z: confounders, X: features in the SML sense, D: treatment in the DML sense. The features X in SML are the conceptual anologous to the treatment D in DML.

7 Comparable to equation (2) for the treatment estimate D, but while D is binary in DML, X here is high-dimensional.

8 Direction-agnostic in the causal sense that it doesn’t tell whether X→Y or Y→X, not in the symmetric sense, as P(Y|X) ≠ P(X|Y).

