## Supplementary Materials for "A causally informed framework for robust confounder control in biomedical machine learning"

*Contents:*

Methods
Supplementary references

### 1. Methods

#### 1.1. Data and pre-processing

For our example predictions we used data of the 1st scanning session (ses-2) of the UK Biobank[1], recorded at three different sites in the UK (Cheadle, Reading, Newcastle). The exact protocol and acquisition parameters of the structural imaging can be found in Miller et al. (2016)[2]. The structural pre-processing was carried out by pipelines developed and run by the UKB[3].

For the Grey Matter Volume (GMV) features 41,180 T1-weighted pre-processed images were retrieved from UKB and converted into a DataLad[4] dataset for provenance tracking with subsequent computations of voxel-based morphometry (CAT 12.7 (default settings); MNI152 space; 1.5mm isotropic)[5]. We extracted the parcel-wise GMV as the winsorized mean (limits 10%) of the voxel-wise values per parcel using the cortical Schaefer et al. (2018)[6] atlas (1000 ROIs), subcortical Tian et al. (2020)[7] (S4 3T) and cerebellar Diedrichsen et al. (2009)[8] (SUIT space) atlas.

All non-imaging variables, including the exemplarily target Hand Grip Strength (HGS) and the investigated example confounders were obtained directly from the UK Biobank[9]. We chose HGS as a robust, objective and reliable target[10–13] to avoid further conceptual problems oftentimes coming along with more latent variables as targets, such as intelligence or executive functioning measures[14]. Healthy subjects were (rather conservatively) defined by excluding the ICD-10 criteria chapters F, G and I60 to I69, which excludes subjects with a history of mental and behavioural disorders, diseases of the nervous system or with a cerebrovascular disease. All NaN values and outliers larger than the 4th standard deviation were removed from the non-imaging data. Additionally, the HGS was averaged over left and right hand and there was a check for balance of sex distributions in the HGS.

#### 1.2. Modelling

10% of the data were set apart to be used as a locked test set for a related project and left untouched for this project. The remaining 90% of the data were split into a training (0.8) and test (0.2) set. Learning algorithms were fitted on the training set by using a cross-validation (CV) scheme. The CV on the training set served to control for the fitting behaviour (e.g. overfitting) of the model and to get an impression of the generalization error. A final estimator, retrained on the entire training set (using root mean squared error; RMSE) was eventually used to make the predictions on the initially held-out test set. These predictions were used to report and visualize the predictive performances. All applied splits were stratified for binned *age*, binned HGS (2 bins) and *sex* (as either defined in the NHS central registry or self-reported). Within the CV scheme, continuous features were z-scored (mean of zero and unit variance). We used a (stratified) 5-fold strategy with one repetition for the CV. We scored the CV using RMSE, mean absolute error (MAE), coefficient of determination ($R^2$), Pearson's r and Spearman's r. The confound removal was applied within the CV to avoid data leakage. Therein, for each feature, a linear regression was fit using the confounds as independent variables and the features as dependent variables. The new, confound-free features were calculated as the residuals of the fitted linear regression (original features minus predicted/fitted features).

#### 1.3. Algorithms, sample sizes and statistical evaluation

The example predictions of HGS from GMV with no confounder adjustment (*vanilla* model) or adjustment for *muscle mass* and *sex* as confounders as illustrated in Fig. 1b (main manuscript) was performed using scikit-learn's[15] linear support vector regression (SVR) with a squared epsilon insensitive loss (L2) and a heuristically calculated hyperparameter C ($C = \frac{1}{\frac{1}{n}\sum_{i=1}^{n}\sqrt{features^2}}$, [16]). This heuristic C value was calculated in a CV consistent manner, i.e. it was calculated only on the training data within the respective fold of the CV. Due to using a heuristic estimate of the hyperparameter C, no nested CV setup was necessary for hyper parameter optimization.

To have comparable models between the unadjusted *vanilla* model and the confounder adjusted model, sample sizes were matched to the variable with the least amount of subjects measured at ses-2, which was *muscle mass* (operationalized through UKB's variable *total lean mass*). This resulted in the shown out-of-sample predictions being performed on N=3620 ($N_{train}$ = 2606, $N_{test}$ = 652) subjects. The 5-fold CV was previously performed on the $N_{train}$ = 2606 subjects.

All correlations were calculated on these same N=3620 subjects. Parcel-wise correlations between parcellated GMV and HGS, parcellated GMV and *muscle mass* as well as HGS and *muscle mass* (all continuous variables) were calculated using Pearson's r (Fig. 1c, Fig. 2 bottom, main manuscript). All correlations including *sex* were calculated using point-biserial correlation coefficient to account for the discrete nature of this variable.

##### 1.4. Code availability

Custom code generated for this project was made publicly available in a GitHub repository. The repository contains further detailed information on used python packages (and versions), code execution and necessary steps for replication of computations.
